# A national study of self-reported COVID symptoms during the first viral wave in Canada

**DOI:** 10.1101/2020.10.02.20205930

**Authors:** Xuyang Tang, Hellen Gelband, Teresa Lam, Nico Nagelkerke, Angus Reid, Prabhat Jha, for the Action to beat Coronavirus (Ab-C) Study Investigators

## Abstract

**Importance:** Accurate understanding of COVID pandemic during the first viral wave in Canada could help prepare for future epidemic waves.

**Objective:** To track the early course of the pandemic by examining self-reported COVID symptoms over time before testing became widely available.

**Design:** Adults from the nationally representative Angus Reid Forum were randomly invited to complete an online survey in May/June 2020. The study is a part of the Action to Beat Coronavirus antibody testing study.

**Setting:** A 20-item internet survey.

**Participants:** 14,408 adults age 18 years of age.

**Exposures:** The months that respondents and any household members first experienced various respiratory, neurological, sleep, skin or gastric symptoms.

**Main Outcomes and Measure:** “COVID symptom-positive,” defined as fever (or fever with hallucinations) plus at least one of difficulty breathing, a dry severe cough, loss of smell or “COVID toe.”

**Results:** In total, 14,408 panel members (48% male and 52% female) completed the survey. Despite overrepresentation of higher levels of education, the prevalence of obesity, smoking, diabetes and hypertension were similar to national census and health surveys. A total of 811 (5.6%) were COVID symptom-positive; highest rates were at ages 18-44 years (8.3% among), declining at older ages. Females had higher odds of reporting COVID symptoms (OR = 1.32, 95%CI 1.11 – 1.56) as did visible minorities (OR = 1.74, 1.29 – 2.35). COVID symptom positivity for respondents and their household members peaked in March (OR = 1.93, 95% CI = 1.59 – 2.34 compared to earlier months).

**Conclusions and Relevance:** This study enhances our current understanding of the progression of the COVID epidemic in Canada, with few laboratory-confirmed cases in January and February, peaking in April. The results suggest substantial viral transmission in March, before widespread testing began, and a gradual decline in cases since May.

## Introduction

The SARS-CoV-2/COVID pandemic has caused serious morbidity and claimed many lives, notably among the elderly, those suffering from chronic illnesses (e.g., diabetes, hypertension, vascular diseases, obesity), or a history of smoking, especially in ethnic minorities.^1^ Canada reported, as of September 1, about 130,000 cases and over 9,000 deaths, the latter mostly in nursing home residents.^2^ Prior to mid-July, testing was reserved for contacts and those with symptoms or high-risk occupations. Thus, the overall proportion of Canadians testing positive (100,000/32 million adults) was well below 0.5%, likely representing only some fraction of all infections during the first viral wave.

Accurately tracking the development of the pandemic helps to understand transmission, and thereby prepare for future epidemic waves. We sought to track the course of the pandemic in Canada before diagnostic testing was widely available by tracking self-reported symptoms of COVID during the early months of viral spread.

The Ab-C study^3^ is a collaboration of the University of Toronto, Unity Health Toronto, and the Angus Reid Institute,^4^ a Canadian public opinion polling organization. This paper reports on results of the first component of the Ab-C study: a survey of COVID symptoms during March-June among a well-characterized representative sample of Canadian adults. Results of the second component—antibody analyses of self-administered dried blood spots from the survey population—will be reported in the coming months.

## Methods

### Study design and survey description

Participants were members of an established panel of Canadians age 18 and older, called the Angus Reid Forum. Forum members were stratified by age, sex, education and province or territories of residence, and then randomly invited on a rolling basis to participate in the online survey portion of the Ab-C study, during which the respondents could opt in for later antibody testing. Members age 60-85 were oversampled because of their elevated risk of serious COVID morbidity and mortality. However, the sampling frame included very few elderly nursing home residents. The Unity Health Toronto Research Ethics Board approved this study (REB# 20-107).

The online survey assessed concerns about COVID of respondents and their household members, whether and when they had or still have specific symptoms typically associated with COVID, testing for COVID and their intention to seek testing, work in high-risk professions for COVID, visits to medical facilities since the beginning of February 2020, and demographic and health status.

### Measures and variables

#### Symptoms

Several retrospective studies that enrolled over a hundred COVID patients in hospitals in mainland China have identified fever^5–11^and cough (dry or unspecified)^5,7–11^ as commonly associated with the infection. Other symptoms experienced by patients in China include a lack of appetite,^6^ general weakness,^6^ malaise,^10^ myalgia or fatigue,^7,10,11^ diarrhea,^6^ excess sputum production,^7^ shortness of breath^8,10^ and sore throat.^11^ A study that examined 1,420 COVID patients from 18 hospitals in Europe found that headache and loss of smell were present in over 70% of the cases.^12^ A cutaneous manifestation (also known as “COVID toe”), appearing similar to skin rash, was common among 375 COVID patients in Spain.^13^ The online Ab-C survey questions about COVID symptoms drew on these findings.

We asked respondents if they had experienced any of the following symptoms that were not related to a condition or illness that they dealt with chronically: difficulty breathing, fever, dry mild cough, severe dry cough (“keeps you from sleeping”), sore throat, frequent sneezing, loss of sense of smell or taste, fever with hallucinations, unusual or disturbed sleep, loss of appetite, dizziness, and red, purple, pink toes with swelling. For each symptom, the possible responses were “Yes, had this but it went away”, “Yes, I still have this”, and “No, have not had this.” We also asked respondents for the month they first had these symptoms, and if they were still having any of them. We asked the respondents the same questions about their household members’ experience with the same symptoms.

We created separate COVID symptom variables for respondents and for household members. We defined COVID symptom-positive as a combination of fever (or fever with hallucinations) with difficulty breathing, a dry cough, loss of smell or “COVID toe.” We created a categorical variable to indicate if no one in the household, only the respondent, only other household member(s), or both the respondent and household member(s) were COVID symptom-positive.

#### Demographics and health

Respondents reported their birth year and month, sex (male/female/prefer to self-describe), highest level of education (some elementary or high school/high school graduate/some college or trade school/graduated from college or trade school/some university/university undergraduate degree/university graduate degree), height, weight, current smoking status (never smoked/smoked daily or occasionally/used to smoke but quit/don’t know/prefer not to answer), diagnosis of diabetes (yes/no/don’t know/prefer not to answer), and diagnosis of high blood pressure (yes/no/don’t know/prefer not to answer). We calculated respondents’ body mass index (BMI) from self-reported height and weight, and categorized BMI as normal or underweight (BMI < 25 kg/m^2^), overweight (25 kg/m^2^ ≤ BMI < 30 kg/m^2^), and obese (BMI ≥ 30 kg/m^2^).^14^ The online poll has existing information on the respondents’ age in years, ethnicity (including whether they self-identify as a visible minority) and region of residence based on their membership information.

### Statistical analysis

We present the proportion of respondents who were COVID symptom-positive by month of onset and age group, and the distribution of COVID symptom-positivity in households by month of onset and age group. We used simple logits to calculate the likelihood of COVID symptom-positivity by month in the overall sample adjusting for known demographic and health risk factors. We excluded responses from 638 respondents with questionable self-reported height (< 100 or > 221 centimeters) and 1,080 respondents with questionable self-reported weight (< 30 or >200 kilograms) from the logit but not for other analyses that do not require weight and height information. We used Stata SE 13^15^ to conduct our analyses.

## Results

A total of 14,408 of 31,839 invited Forum members completed the online survey. The demographics and health characteristics of those completing the online poll are summarized in Table 1, alongside the proportion of population with those characteristics in the Canadian population overall, from various sources. The distribution of demographic characteristics of the respondents in the Ab-C study are similar to those from the 2016 Census Canada,^16–19^ with the exception of an overrepresentation of adults with some university education (49% in our sample versus 32% in Canada). Despite this, the Ab-C study participants were quite similar to the national prevalences of obesity, current smoking, diabetes and hypertension, as reported by Public Health Agency of Canada,^20^ Health Reports^21^ and Health Canada. ^22,23^

**Table 1.** Demographic, health characteristics and experience of any symptom in the sample with comparison to 2016 Census Canada, Public Health Agency of Canada, Stats Can and Health Canada

| Characteristics | Canada | Ab-C |
| --- | --- | --- |
|  | % | No. (%) |
| Age categories <sup>a</sup> |  |  |
| 18 to 44 | 36% | 5,006 (35%) |
| 45 to 64 | 27% | 5,415 (38%) |
| 65 to 74 | 10% | 3,228 (22%) |
| 75 and older | 7% | 759 (5%) |
| Sex <sup>a</sup> |  |  |
| Male | 49% | 6,911 (48%) |
| Female | 51% | 7,424 (52%) |
| Prefer to self-describe |  | 73 (1%) |
| Education <sup>a</sup> |  |  |
| High school graduate or under | 35% | 2,070 (14%) |
| Some college or college graduate | 33% | 5,316 (37%) |
| Some university or higher | 32% | 7,022 (49%) |
| Household size <sup>a</sup> |  |  |
| Just one – I live alone | 28% | 2,638 (18%) |
| Two people | 34% | 6,417 (45%) |
| Three people | 15% | 2,363 (16%) |
| Four people | 14% | 1,923 (13%) |
| Five or more people in my household | 8% | 1,067 (7%) |
| Respondent ethnicity <sup>a</sup> |  |  |
| Indigenous | 5% | 696 (5%) |
| English and other European | 73% | 12,191 (85%) |
| Others | 22% | 1,140 (8%) |
| Rather not say |  | 381 (3%) |
| Visible Minority <sup>a</sup> |  |  |
| No | 78% | 12,797 (89%) |
| Yes | 22% | 1,611 (11%) |
| Province <sup>a</sup> |  |  |
| Ontario | 38% | 5,601 (39%) |
| British Columbia | 13% | 2,072 (14%) |
| Quebec | 23% | 2,887 (20%) |
| Prairie provinces | 18% | 2,679 (19%) |
| Atlantic provinces | 7% | 1,169 (8%) |
| Weight status (calculated from Body Mass Index) <sup>d</sup> |  |  |
| Under or Normal Weight (BMI < 25 kg/m<sup>2</sup>) | 37% | 4,130 (33%) |
| Overweight (25 kg/m<sup>2</sup> ≤ BMI < 30 kg/m<sup>2</sup>) | 37% | 4,690 (37%) |
| Obese (BMI ≥ 30 kg/m<sup>2</sup>) | 27% | 3,870 (31%) |
| Currently smoking |  |  |
| No |  | 12,465 (86%) |
| Yes (Daily or occasional) <sup>d</sup> | 15% | 1,943 (14%) |
| Ever diagnosed as diabetic |  |  |
| No |  | 12,736 (88%) |
| Yes <sup>b</sup> | 9% | 1,536 (11%) |
| Do not know or Preferred not to answer |  | 136 (1%) |
| Ever diagnosed as hypertensive |  |  |
| No |  | 9,951 (69%) |
| Yes <sup>c</sup> | 23% | 4,241 (29%) |
| Do not know or Preferred not to answer |  | 216 (2%) |
| Experience with <u>any</u> symptom |  |  |
| Had symptom before March |  | 2,198 (15%) |
| Had symptom in March |  | 1,789 (12%) |
| Had symptom in April |  | 1,264 (9%) |
| Had symptom in May or later |  | 962 (7%) |
| Had symptom but cannot recall the month |  | 1,096 (8%) |
| Did not have symptom |  | 7,099 (49%) |

Sources: a. 2016 Census Canada<sup>16-19</sup>; b. Public Health Agency of Canada<sup>20</sup>; c. Health Reports<sup>21</sup>; d. Health Canada<sup>22,23</sup>

### Time trends

In the sample, 7,309 respondents experienced at least one of the survey symptoms, and 811 (6% of the entire cohort) met the study definition of COVID symptom-positivity (Table 2). Table 2 presents respondents’ prevalences and odds ratios (OR) of COVID symptoms. After adjusting for age, sex, ethnicity, education, province of residence, BMI, smoking status, and history of diabetes and hypertension, the likelihood of COVID symptom-positivity were highest among younger adults aged 18-44 (8.3%), and fell in older age groups, to 2.9% at 65-74 years and 3.4% at 75 years and older (OR of about 0.48 in the last two age groups). Females had higher odds of reporting COVID symptoms (OR = 1.32, 95%CI 1.11 – 1.56) as did visible minorities (OR = 1.74, 1.29 – 2.35).

**Table 2.**
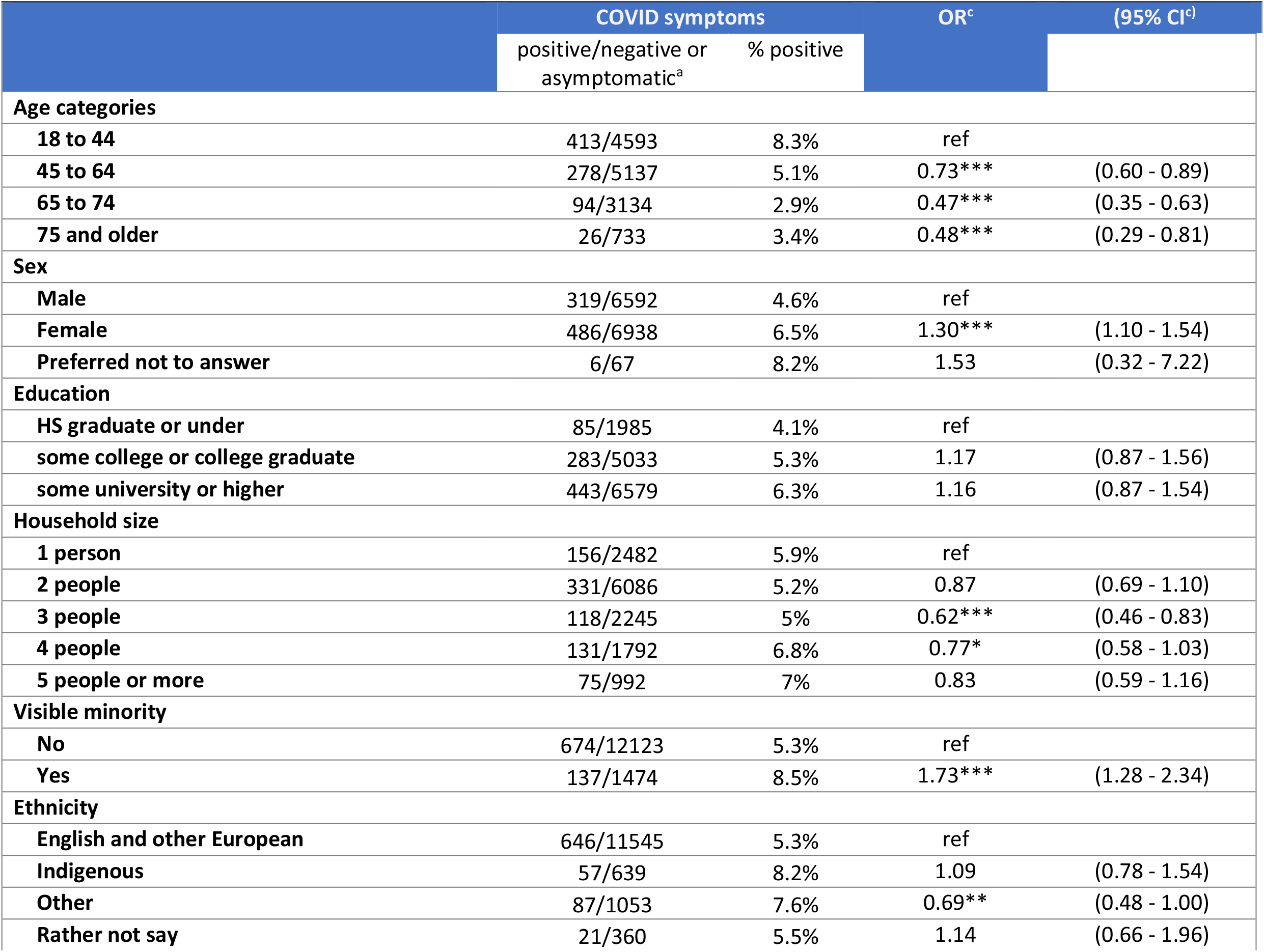
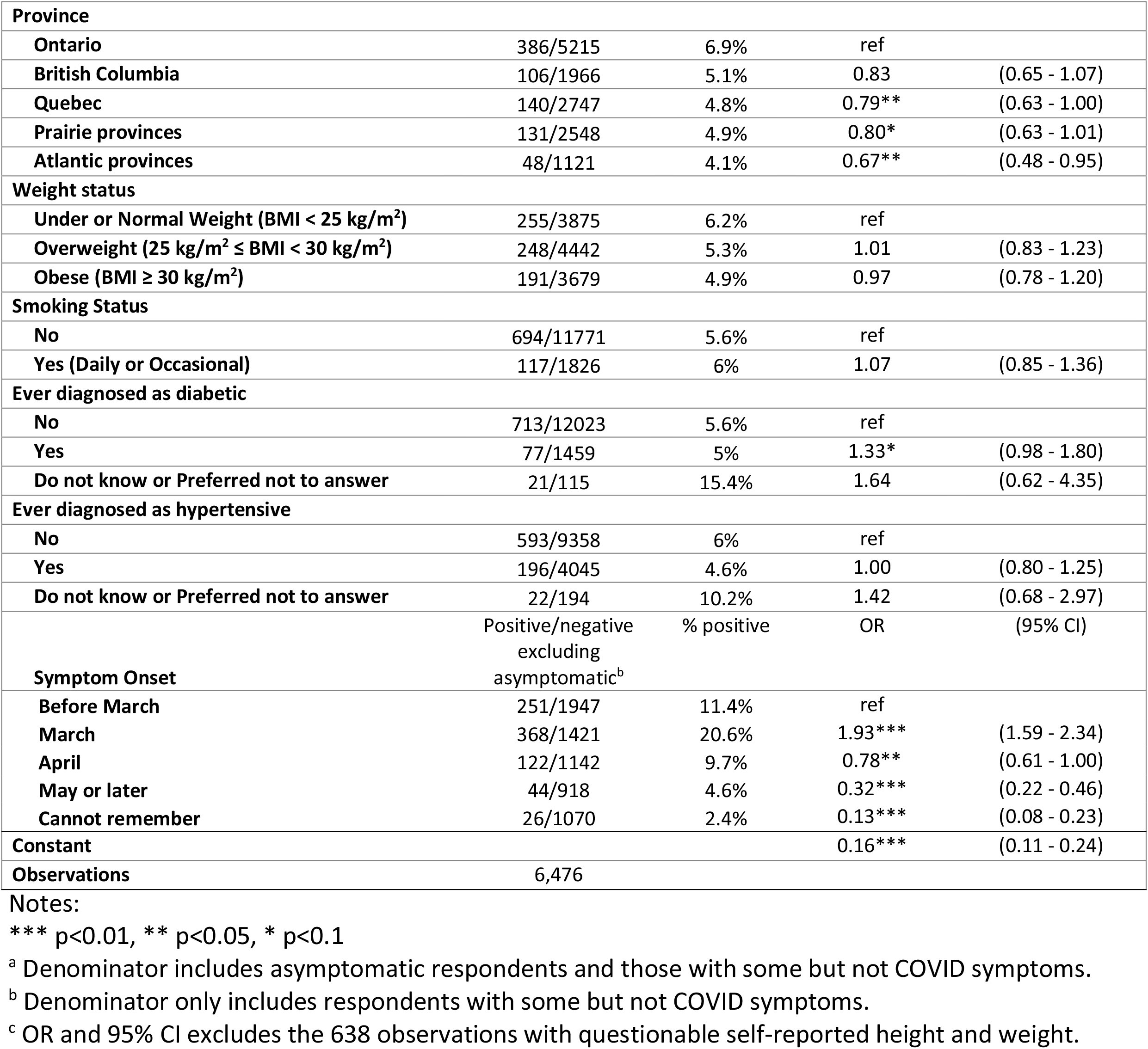
Respondents’ odds ratio of COVID symptom positivity

|  | COVID symptoms |  | OR<sup>c</sup> | (95% CI<sup>d</sup>) |
| --- | --- | --- | --- | --- |
|  | positive/negative or asymptomatic<sup>a</sup> | % positive |  |  |
| Age categories |  |  |  |  |
| 18 to 44 | 413/4593 | 8.3% | ref |  |
| 45 to 64 | 278/5137 | 5.1% | 0.73*** | (0.60 - 0.89) |
| 65 to 74 | 94/3134 | 2.9% | 0.47*** | (0.35 - 0.63) |
| 75 and older | 26/733 | 3.4% | 0.48*** | (0.29 - 0.81) |
| Sex |  |  |  |  |
| Male | 319/6592 | 4.6% | ref |  |
| Female | 486/6938 | 6.5% | 1.30*** | (1.10 - 1.54) |
| Preferred not to answer | 6/67 | 8.2% | 1.53 | (0.32 - 7.22) |
| Education |  |  |  |  |
| HS graduate or under | 85/1985 | 4.1% | ref |  |
| some college or college graduate | 283/5033 | 5.3% | 1.17 | (0.87 - 1.56) |
| some university or higher | 443/6579 | 6.3% | 1.16 | (0.87 - 1.54) |
| Household size |  |  |  |  |
| 1 person | 156/2482 | 5.9% | ref |  |
| 2 people | 331/6086 | 5.2% | 0.87 | (0.69 - 1.10) |
| 3 people | 118/2245 | 5% | 0.62*** | (0.46 - 0.83) |
| 4 people | 131/1792 | 6.8% | 0.77* | (0.58 - 1.03) |
| 5 people or more | 75/992 | 7% | 0.83 | (0.59 - 1.16) |
| Visible minority |  |  |  |  |
| No | 674/12123 | 5.3% | ref |  |
| Yes | 137/1474 | 8.5% | 1.73*** | (1.28 - 2.34) |
| Ethnicity |  |  |  |  |
| English and other European | 646/11545 | 5.3% | ref |  |
| Indigenous | 57/639 | 8.2% | 1.09 | (0.78 - 1.54) |
| Other | 87/1053 | 7.6% | 0.69** | (0.48 - 1.00) |
| Rather not say | 21/360 | 5.5% | 1.14 | (0.66 - 1.96) |
| **Province** |  |  |  |  |
| Ontario | 386/5215 | 6.9% | ref |  |
| British Columbia | 106/1966 | 5.1% | 0.83 | (0.65 - 1.07) |
| Quebec | 140/2747 | 4.8% | 0.79** | (0.63 - 1.00) |
| Prairie provinces | 131/2548 | 4.9% | 0.80* | (0.63 - 1.01) |
| Atlantic provinces | 48/1121 | 4.1% | 0.67** | (0.48 - 0.95) |
| **Weight status** |  |  |  |  |
| Under or Normal Weight (BMI < 25 kg/m<sup>2</sup>) | 255/3875 | 6.2% | ref |  |
| Overweight (25 kg/m<sup>2</sup> ≤ BMI < 30 kg/m<sup>2</sup>) | 248/4442 | 5.3% | 1.01 | (0.83 - 1.23) |
| Obese (BMI ≥ 30 kg/m<sup>2</sup>) | 191/3679 | 4.9% | 0.97 | (0.78 - 1.20) |
| **Smoking Status** |  |  |  |  |
| No | 694/11771 | 5.6% | ref |  |
| Yes (Daily or Occasional) | 117/1826 | 6% | 1.07 | (0.85 - 1.36) |
| **Ever diagnosed as diabetic** |  |  |  |  |
| No | 713/12023 | 5.6% | ref |  |
| Yes | 77/1459 | 5% | 1.33* | (0.98 - 1.80) |
| Do not know or Preferred not to answer | 21/115 | 15.4% | 1.64 | (0.62 - 4.35) |
| **Ever diagnosed as hypertensive** |  |  |  |  |
| No | 593/9358 | 6% | ref |  |
| Yes | 196/4045 | 4.6% | 1.00 | (0.80 - 1.25) |
| Do not know or Preferred not to answer | 22/194 | 10.2% | 1.42 | (0.68 - 2.97) |
| **Symptom Onset** |  |  |  |  |
|  | Positive/negative<br>excluding<br>asymptomatic<sup>b</sup> | % positive | OR | (95% CI) |
| Before March | 251/1947 | 11.4% | ref |  |
| March | 368/1421 | 20.6% | 1.93*** | (1.59 - 2.34) |
| April | 122/1142 | 9.7% | 0.78** | (0.61 - 1.00) |
| May or later | 44/918 | 4.6% | 0.32*** | (0.22 - 0.46) |
| Cannot remember | 26/1070 | 2.4% | 0.13*** | (0.08 - 0.23) |
| Constant |  |  | 0.16*** | (0.11 - 0.24) |
| Observations | 6,476 |  |  |  |

Notes:
\*\*\* $p < 0.01$ , \*\* $p < 0.05$ , \* $p < 0.1$
<sup>a</sup> Denominator includes asymptomatic respondents and those with some but not COVID symptoms.
<sup>b</sup> Denominator only includes respondents with some but not COVID symptoms.
<sup>c</sup> OR and 95% CI excludes the 638 observations with questionable self-reported height and weight.

Figure 1 shows the proportion of respondents COVID symptom-positive in each month among those reporting any symptom in all ages and by age group. March had the highest proportion of COVID symptom-positive respondents (20.6%) of those reporting any symptom in all age groups. In April, a higher proportion of respondents age 18-44 were COVID symptom-positive (12.2%) than respondents in older age groups (9.3% in age 45-64, 4.2% in age 65-74 and 2.6% in age 75 years and older). However, the 75+ group saw an increase (8.6%) of COVID symptom-positivity in May or later. The corresponding OR compared to February or earlier were in March (OR = 1.93, 1.59 – 2.34), falling in April (OR = 0.78, 0.61 – 1.00) and May or later (OR = 0.32, 0.08 – 0.23). (Table 2) The counts of COVID symptom positivity among those who experienced any symptom by month and age group are presented in Appendices A and B.

**Figure 1.**
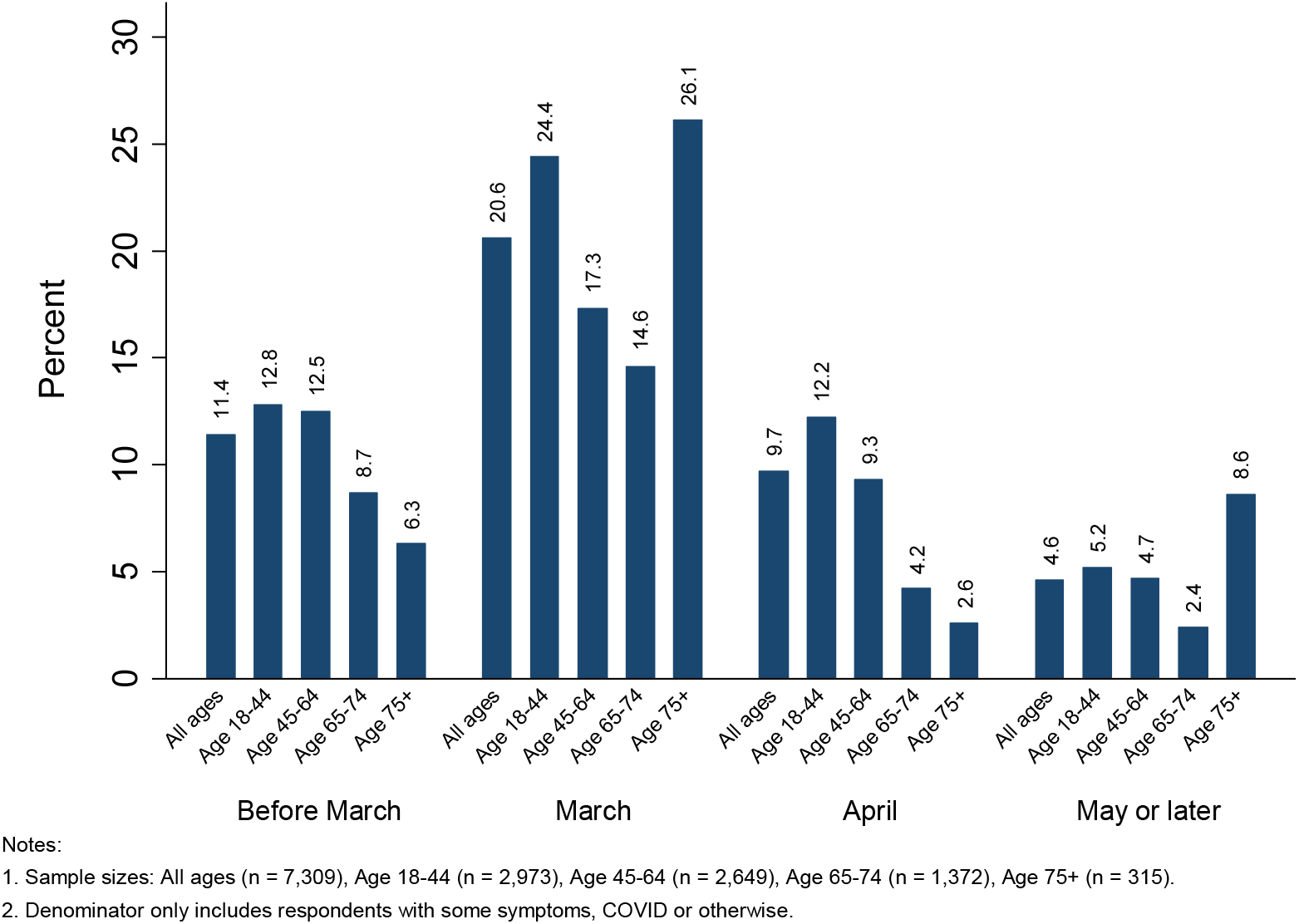
COVID symptom positivity by month of onset and age group.

In the sample with at least two household members, the proportion of those reporting COVID symptoms was the same (5.6%). In these households, 4,175 reported any symptom in someone else aside from the respondent of which 651 (16%) reported COVID symptoms. Figure 2 presents the distribution of COVID symptom positivity in households with at least two members, by month and respondents’ age group. Of the 651 households, 351 reported COVID symptom only in household members but not the respondent, while 300 reported COVID symptom in both household members and the respondent. March was also the peak of COVID symptom positivity for respondents and/or their household members for all age groups, followed by declines of COVID symptom positivity in April and later months. In March, respondents aged 75 and above were more likely than younger respondents to be the only one in a multi-person household with symptoms (21% in age 75+ vs. 11% in age 18-44, 8% in age 45-64, 10% in age 65-74).

**Figure 2.**
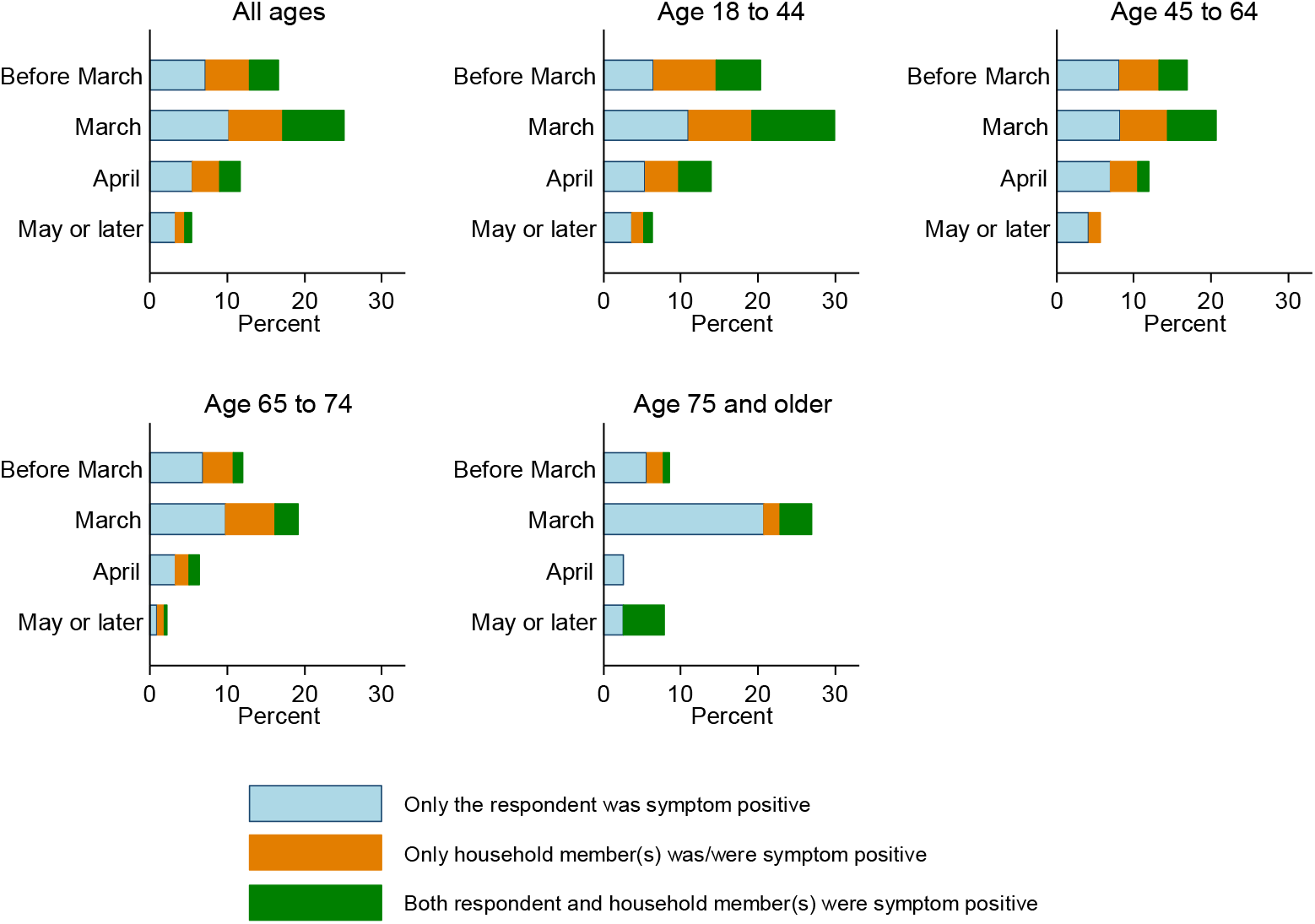
Household distribution of COVID symptom by time and respondent age group.

## Discussion

In our sample of 14,408 Canadian adults, 5.6% of met the study definition of COVID symptom-positivity during the March-June period of Canada’s first viral wave. The highest prevalence was among younger adults, women and visible minorities. COVID symptoms were highest in March 2020, at 2.6%. Results were similar among household respondents, as has been noted for an earlier smaller national poll^24^. Among our respondents, most of the demographic and health risk factors for COVID mortality established in earlier studies (e.g., smoking status, obesity, history of diabetes or high blood pressure)^1,25,28^ were not associated with elevated odds of COVID symptom-positivity, suggesting that such factors may not affect the likelihood of developing symptoms but rather increase the probability of hospitalization and dying from COVID once infected or developing symptoms.

The March peak of symptoms is roughly consistent with the current understanding of the pandemic in Canada. Until about mid-July, only symptomatic patients, close contacts, those with a travel history to high-risk regions or high-risk occupations were permitted SARS-COV-2 testing. The official peak, based on testing, was April, but it is likely based on our results that significant transmission was taking place earlier. This is also consistent with the May peak in COVID deaths^26^ (Figure 3). Most of the COVID deaths occurred in nursing home populations, but community deaths followed a similar pattern.^2^

**Figure 3.**
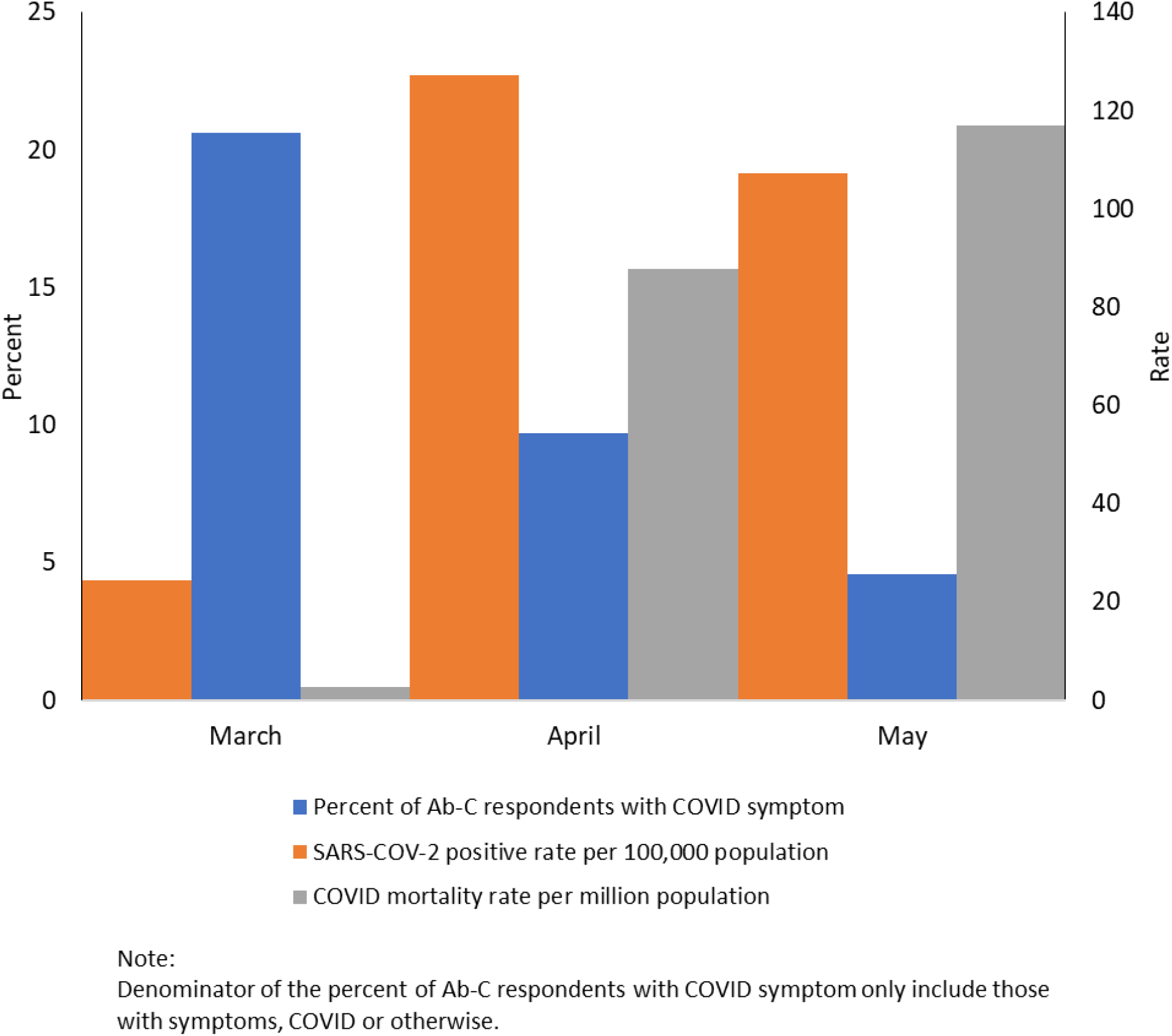
COVID symptom positivity among Ab-C respondents compared to COVID cases and deaths in Canada.

A strength of this study is using a sample with demographics and health characteristics similar to that of all Canadian adults from a well-characterized polling panel. The study has certain limitations. Symptom positivity data are available only monthly rather than weekly, which would have given us a finer tuned look at the shape of the symptom curve. However, monthly data are less likely to be misreported retrospectively. Also, there is the potential for reporting biases. Overreporting, i.e., low specificity of symptoms for COVID, could result from time-varying knowledge and awareness of COVID effects. We examined household distribution of less common symptoms (e.g., loss of appetite, dizziness, disturbed sleep) to see whether participants and their household members simply noticed symptoms because of the pandemic or whether they truly experienced symptoms, using graphs similar to Figure 2. The shape of the graphs is generally consistent with Figure 2 which suggests that respondents may not have enhanced reporting of symptoms as they become more aware of COVID (data not shown). Naturally, self-reported symptoms may represent not just COVID but other conditions, including seasonal influenza. Symptoms also have low sensitivity for COVID as notable proportion of (infectious) COVID cases are asymptomatic.^27^

The next phase of the study will as combine survey responses with antibody testing results as well as examining the discordance of symptoms and antibodies in multi-person households. The antibody collection started in mid-June, which should allow for testing of at least IgG antibodies which have been shown to be stable for a few months after infection^28^. As such, it will provide a unique insight on the first viral wave in Canada.

This study enhances our current understanding of the progression of the COVID epidemic in Canada, with symptoms peaking in March. There were few laboratory-confirmed cases in January and February, with these peaking in April. The results suggest substantial viral transmission in March, before widespread testing began, and a gradual decline in cases since May.^29^

## Data Availability

The data that support the findings of the Ab-C Study are available on request from the corresponding author PJ. The request is also subject to the approval of the Unity Health Toronto Research Ethics Board.

### Appendices

#### Appendix A

Counts of COVID symptom in respondents and household members by age groups and month of symptoms’ onset.

Part A. Number of respondents with COVID symptom among those reported any symptom by the month of symptoms’ onset and age group

| Age group | COVID symptom | Before March | March | April | May or later | Cannot recall | Total |
| --- | --- | --- | --- | --- | --- | --- | --- |
| All ages | No | 1,947 | 1,421 | 1,142 | 918 | 1,070 | 6,498 |
|  | Yes | 251 | 368 | 122 | 44 | 26 | 811 |
| Age 18 to 44 | No | 653 | 651 | 503 | 381 | 372 | 2,560 |
|  | Yes | 96 | 210 | 70 | 21 | 16 | 413 |
| Age 45 to 64 | No | 732 | 525 | 418 | 305 | 391 | 2,371 |
|  | Yes | 105 | 110 | 43 | 15 | 5 | 278 |
| Age 65 to 74 | No | 443 | 211 | 183 | 200 | 241 | 1,278 |
|  | Yes | 42 | 36 | 8 | 5 | 3 | 94 |
| Age 75 and older | No | 119 | 34 | 38 | 32 | 66 | 289 |
|  | Yes | 8 | 12 | 1 | 3 | 2 | 26 |

Part B. Number of household members (in 2+ member households) with COVID symptom among those reported any symptom by the month of symptoms’ onset and age group

| Age group | COVID symptom | Before March | March | April | May or later | Cannot recall | Total |
| --- | --- | --- | --- | --- | --- | --- | --- |
| All ages | No | 789 | 897 | 687 | 561 | 590 | 3,524 |
|  | Yes | 176 | 314 | 98 | 46 | 17 | 651 |
| Age 18 to 44 | No | 280 | 453 | 347 | 276 | 236 | 1,592 |
|  | Yes | 89 | 195 | 50 | 29 | 12 | 375 |
| Age 45 to 64 | No | 297 | 336 | 237 | 190 | 202 | 1,262 |
|  | Yes | 63 | 91 | 34 | 11 | 4 | 203 |
| Age 65 to 74 | No | 168 | 95 | 91 | 83 | 119 | 556 |
|  | Yes | 21 | 26 | 12 | 4 | 1 | 64 |
| Age 75 and older | No | 44 | 13 | 12 | 12 | 33 | 114 |
|  | Yes | 3 | 2 | 2 | 2 | 0 | 9 |

#### Appendix B

Ab-C Study Investigators

##### St. Michael’s Hospital (Unity Health Toronto)

*Centre for Global Health Research* P Jha (Principal Investigator), P Brown (scientist), H Gelband (senior fellow), N Nagelkerke (senior fellow), E Young (coordinator), A Sharma (coordinator), P Rodriguez (coordinator), C Schultz (coordinator), C Birnboim (senior consultant), L Newcombe; *Applied Health Research Centre*, P Juni (scientist); *Keenan Research Centre for Biomedical Science* H Ni (platform director, Scientist); *Immunology Laboratory* M Pasic (Division Head); *Li Ka Shing Knowledge Institute* G Booth (scientist); Arthur Slutsky, Scientist (scientist, Chair, Scientific Advisory Board)

##### University Health Network

I Bogoch (scientist)

##### Sinai Health

AC Gingras (scientist), K Colwill (coordinator)

##### Children’s Hospital of Eastern Ontario and University of Ottawa

P Chakraborty (scientist), MA Langlois (scientist)

##### Angus Reid Forum

A Reid, E Morawski, D Eliopoulos, A Hollander

